# Effectiveness of Osteopathic Manipulative Treatment for Structural Musculoskeletal Pain: A Meta-Analysis of Randomized Controlled Trials

**DOI:** 10.64898/2026.08.12.26359899

**Authors:** Anjolina L. Hsiao, Garrett C. Schimmel, Rohan U. Kale, Miguel G. Dimanlig, Manoela Ortegosa da Cunha, Nicole Myers

## Abstract

Structural musculoskeletal pain, defined as pain associated with musculoskeletal conditions of the spine and peripheral joints, afflicts persons widely, independent of demographic, and continues to contribute substantially to disability on a global scale.

Osteopathic manipulative treatment (OMT) is a non-invasive therapy performed by osteopathic physicians, encompassing a wide variety of techniques meant to heal the dysfunctions manifesting structural musculoskeletal pain. However, the efficacy of OMT in relieving pain symptomatology remains subject to debate.

This meta-analysis examines the effect OMT serves to manage structural musculoskeletal pain, measured on a Visual Analog Scale. Three randomized control studies (RCTs) were included, with a total of 231 participants, 117 of which received OMT as part of pain management treatment, the other 114 receiving other treatment modalities.

Using the random effects model, the mean difference between OMT and non-OMT treated groups was -1.80 (−7.31; 3.78). Although this mean difference favors OMT with regard to greater reduction in pain, the finding is not statistically significant. Heterogeneity was found to be extraordinarily high (I^2^ = 96%) and statistically significant (p = <0.0001), albeit attributed to one of the papers, deemed an outlier. With its removal, heterogeneity was still moderate (I^2^ = 54.4%). Given these findings, the efficacy of OMT in reducing structural musculoskeletal pain cannot be proven.

A significant limitation of this study was a low sample size, consisting of 3 RCTs, reducing statistical power. In addition, there was high heterogeneity between studies. More high-quality RCTs with larger sample sizes, standardized methods, and an examination of a broader set of structural musculoskeletal conditions are necessitated to better evaluate the contribution of OMT in pain reduction.

## Introduction

Musculoskeletal pain is a major contributor to disability worldwide and can substantially impair mobility, activities of daily living, occupational participation, and quality of life. It encompasses a heterogeneous group of conditions involving the spine and peripheral joints, with clinical manifestations and treatment requirements that vary according to the underlying diagnosis. For the purposes of the present meta-analysis, structural musculoskeletal pain is defined operationally as pain associated with musculoskeletal conditions of the spine or peripheral joints represented in the eligible literature, including knee osteoarthritis, shoulder pain, and cervical spondylosis^1^.

Management of musculoskeletal pain is generally individualized and multimodal. Depending on the underlying condition and its severity, treatment may include patient education, therapeutic exercise and rehabilitation, pharmacologic therapy, injections, and surgical intervention. Because persistent pain may remain despite standard management and no single intervention is uniformly effective across musculoskeletal diagnoses, nonpharmacologic approaches are frequently incorporated into comprehensive treatment plans. Osteopathic manipulative treatment (OMT) is a hands-on component of osteopathic care that may be used to complement, rather than replace, conventional medical management^2^.

Evidence regarding the effectiveness of OMT for musculoskeletal pain remains mixed^1^. An overview of systematic reviews reported possible benefits for selected musculoskeletal disorders, particularly nonspecific low-back and neck pain; however, the methodological quality of the included reviews was rated as low or critically low, and the literature was limited by small samples, conflicting findings, and heterogeneity among treatment protocols. More recent findings have also been inconsistent. A 2024 meta-analysis restricted to sham- or placebo-controlled trials did not demonstrate that OMT was superior to sham or placebo for improving pain, disability, or quality of life in patients with neck or low-back pain. Similarly, a 2025 meta-analysis of localized joint pain found differing conclusions between the common-effect and random-effects models in the presence of substantial heterogeneity, preventing a firm conclusion regarding overall efficacy^1^.

Randomized controlled trials have examined osteopathic manual interventions in patients with knee osteoarthritis, shoulder pain, and cervical spondylosis. However, differences in patient populations, intervention protocols, comparator groups, and pain-assessment methods complicate synthesis and limit the generalizability of individual study findings. Therefore, the objective of this meta-analysis was to evaluate the effect of OMT-containing care on pain intensity in adults with structural musculoskeletal pain by pooling eligible randomized controlled trials comparing OMT-containing interventions with non-OMT comparators. We hypothesized that OMT- containing care would produce a greater reduction in pain intensity than non-OMT care.

## Material Methods

### 2.1 Procedures

The Preferred Reporting Items for Systematic Reviews and Meta-Analyses (PRISMA) criteria was followed for this review^3^. The research question studied is defined as: Is there a significant difference in pain reduction when OMT is used to manage structural pain than non- OMT management?

Inclusion of studies in this meta analysis was based on meeting eligibility criteria. All studies included must have compared an OMT treated group to a non-OMT treated group in patients with structural pain. Studies must have reported pain intensity following treatment on the Visual Analog Scale (VAS) scale.

Excluded studies include meta analysis, cross-sectional studies, prospective and retrospective cohort studies, observational studies, case reports and all other non-RCTs. All studies published before 2016 were excluded from analysis. All studies that did not report pain on a VAS scale were excluded. All studies that did not pertain to OMT or structural pain in regard to its definition were excluded.

Pubmed and Google Scholar were scoured for all RCTs studying the effect of OMT on pain management through July 2026. Search terms included “OMT”, “Osteopathic manipulative treatment”, “pain management”, “pain”, “arthritis”, “spondylosis”, “bursitis”, “fracture”, “RCT”, and “Randomized Control Trial”. This resulted in a culmination of 157 studies, which were then screened through based on eligibility (inclusion/exclusion criteria) to be used in the study. A total of 157 articles were screened for eligibility. 29 records were identified as duplicates and removed. 120 articles were screened out for being non-RCTs, published before 2016, or because the pain did not meet the definition of being structural. After screening, 8 articles were left to be included in the meta analysis. 3 were removed due to pain not being reported on the VAS scale, and 2 were removed due to both groups in the studies receiving OMT treatment in comparison to one another.

Data pertaining to title, author, year, sample size, study design, mean pain on VAS, standard deviation, were extracted from each study.

Only RCTs were included in this study to minimize bias risk. Bias assessment was conducted using the Cochrane Risk of Bias Tool^4^ and the National Institute of Health (NIH) quality assessment tool^5^.

### 2.2 Data analysis

Data analysis was performed by hand with pen and paper and repeated in excel. Forest plots were generated using Graphpad Prism (v11.0.2). Forest plots were generated regarding all three studies for pain on VAS and excluding the outlier study (Schwerla et al. 2024^6^) with each study’s mean value, standard deviation, and associated confidence interval. Schwerla et al. 2020^6^ had a VAS score on a scale of 0-100 and was scaled down to match the 0-10 model of the other two studies. A random-effects model, with 95% confidence intervals, was calculated. Heterogeneity was assessed using the I² and Tau² statistics to evaluate variation across studies.

Funnel plots were generated using RStudio (v4.5.1, R Core Team, 2025) and the meta package (v7.1-0). Funnel plots were generated displaying publication bias in regard to pain reduction of OMT groups and non OMT groups treated for structural pain of all three studies and excluding the outlier study (Schwerla et al. 2020^6^). Funnel plots were created using mean difference against standard error of the three included studies.

Forest plots that were created for this meta-analysis include calculations for both the random-effects and common-effects models. Further sections will analyze the random effects model. The random effects model is the most appropriate structure to interpret the study results.

## Results

Following our literature search, 157 articles were identified. After screening to meet all inclusion and exclusion criteria, three studies were included in our meta-analysis (Figure 1).

**Figure 1:**
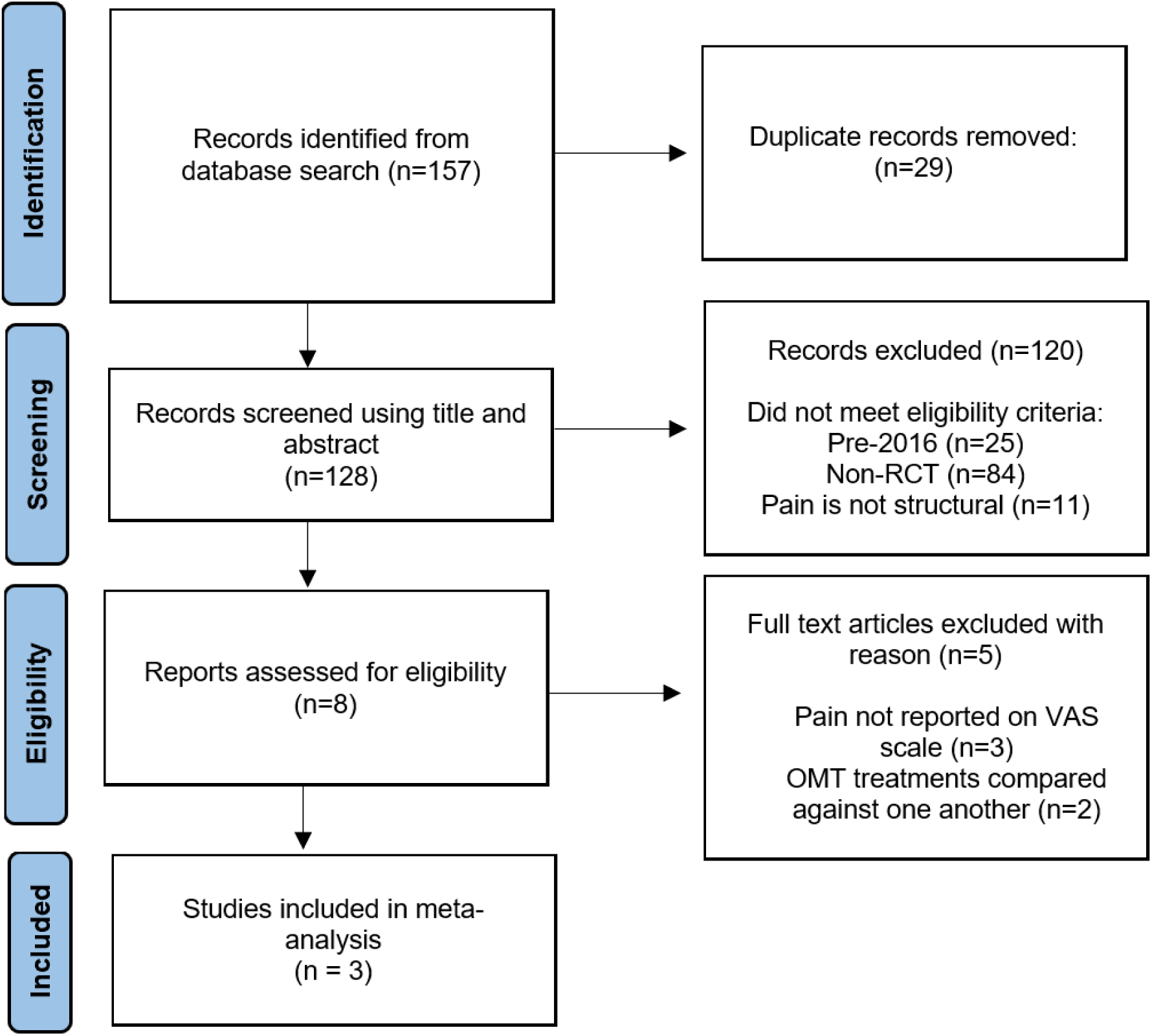
PRISMA flow chart for literature search and selection process. n: number; OMT: osteopathic manipulative treatment; non-RCT: non-randomized controlled trial; VAS: visual analog scale; PRISMA: Preferred Reporting Items for Systematic Reviews and Meta-Analyses

[ufig1]

Study characteristics of the three studies included in our meta-analysis are summarized in Table 1.

**Table 1:** Summary of the study characteristics included in the data analysis.

| Study Name | Total Sample Size (n) | OMT Group Participants (n) | Control Group Participants (n) | Study Design | OMT treatment used | Outcomes measured |
| --- | --- | --- | --- | --- | --- | --- |
| Altinbilek et al | 85 | 43 | 42 | RCT | Joint mobilization/ compression, lymphatic drainage | Pain on VAS scale, ROM |
| Schwerla et al | 70 | 36 | 34 | RCT | Treatment depended on patient assessment | Pain on VAS scale, disability, pain frequency |
| Sezerel et al | 76 | 38 | 38 | RCT | Muscle Energy Technique | Pain on VAS scale, disability, neck positional sense |

n: number; OMT: osteopathic manipulative treatment; RCT: randomized controlled trial; VAS: visual analog scale; ROM: range of motion

| Study Name | Total Sample Size (n) | OMT Group Participants (n) | Control Group Participants (n) | Study Design | OMT treatment used | Outcomes measured |
| --- | --- | --- | --- | --- | --- | --- |
| Altinbilek et al. 2018<sup>7</sup> | 85 | 43 | 42 | RCT | Joint mobilization/ compression, lymphatic drainage | Pain on VAS scale, ROM |
| Schwerla et al. 2020<sup>6</sup> | 70 | 36 | 34 | RCT | Treatment depended on patient assessment | Pain on VAS scale, disability, pain frequency |
| Sezerel et al. 2024<sup>8</sup> | 76 | 38 | 38 | RCT | Muscle Energy Technique | Pain on VAS scale, disability, neck positional sense |

Risk of bias assessment was conducted originally using the Cochrane Risk of Bias Tool^4^. The Cochrane Risk of Bias Assessment was conducted independently by two separate reviewers (A.H. and G.S.). No discrepancies or disagreements were found between reviewers. Bias assessment was conducted by completing the guidance document for each included study, with each of five domains being scored from low risk to high risk. The domains were then tallied and each paper was determined to be of low risk to high risk of bias. Results from this bias assessment can be found in Table 2. The bias ruling for Altinbilek et al. 2018^7^ was “high risk”, the bias ruling for Schwerla et al. 2020^6^ was “high risk’, and the bias ruling for Sezerel et al. 2024^8^, was “high risk.” A note was made that high risk determinations were largely driven by Domain 2 (Blinding). This is an intrinsic, unavoidable limitation of manual medicine research, rather than a failure of the investigators’ trial design.

**Table 2:** Risk-of-Bias Assessment for Included Studies in Accordance to the Cochrane Risk of Bias Tool.

| Study | D1 | D2 | D3 | D4 | D5 | Overall Bias |
| --- | --- | --- | --- | --- | --- | --- |
| Altinbilek et al. | Low | High | Low | Concerns | Low | High Risk |
| Schwerla et al. | Low | High | Low | Concerns | Low | High Risk |
| Sezerel et al. | Low | High | Low | Concerns | Low | High Risk |

D1-D5 represent the 5 domains used in the Cochrane Risk of Bias Tool. Studies are rated “low risk”, “concerns”, “high risk” for each of the 5 domains.
D1: domain 1 (randomization); D2: domain 2 (deviations); D3: domain 3 (missing data), D4: domain 4 (measurement); D5: domain 5 (reporting)

| Study | D1 | D2 | D3 | D4 | D5 | Overall Bias |
| --- | --- | --- | --- | --- | --- | --- |
| Altinbilek et al. 2018<sup>7</sup> | Low | High | Low | Concerns | Low | High Risk |
| Schwerla et al. 2020<sup>6</sup> | Low | High | Low | Concerns | Low | High Risk |
| Sezerel et al. 2024<sup>8</sup> | Low | High | Low | Concerns | Low | High Risk |

Risk of bias assessment was expanded upon using the NIH Study Quality Assessment Tool^5^. The Quality Assessment Tool was conducted independently by two separate reviewers (A.H. and G.S.). No discrepancies or disagreements were found between reviewers. Scoring was conducted by awarding 0 points for “no” responses and 1 point for “yes” responses to the quality assessment tool questions. A score of 0-4 points equated to “poor”, a score of 5-9 points equated to “fair”, and a score of 10-14 points equated to “good.” Results from the quality assessment can be found in Table 3. The bias ruling for Altinbilek et al. 2018^7^ was “fair”, the bias ruling for Schwerla et al. 2020^6^ was “fair’, and the bias ruling for Sezerel et al. 2024^8^, was “fair.” Funnel plots were employed to display risk of publication bias of each included study.

**Table 3:**
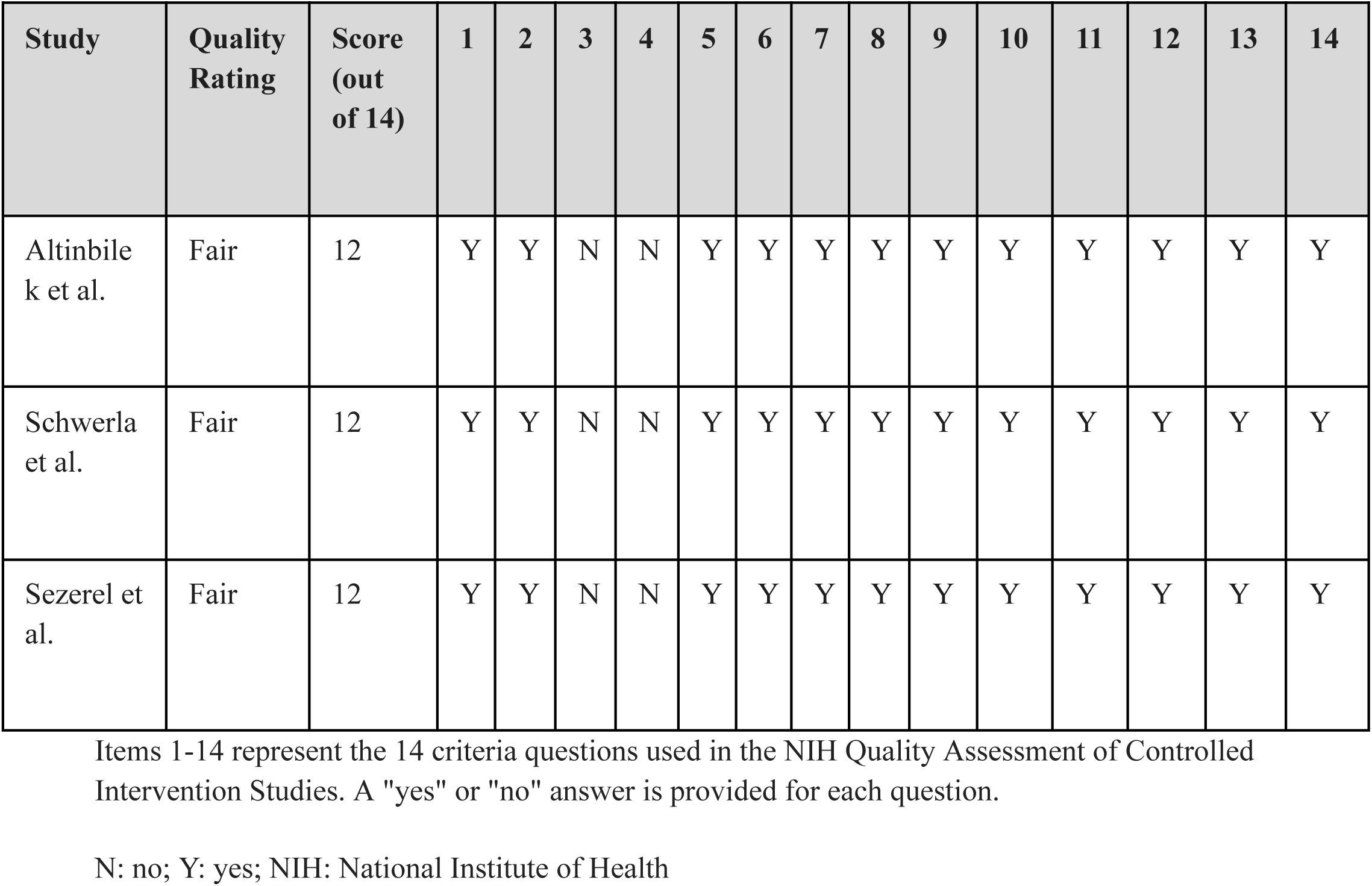
Risk-of-Bias Assessment for Included Studies in Accordance to the NIH Quality Assessment Tool.

| Study | Quality Rating | Score (out of 14) | 1 | 2 | 3 | 4 | 5 | 6 | 7 | 8 | 9 | 10 | 11 | 12 | 13 | 14 |
| --- | --- | --- | --- | --- | --- | --- | --- | --- | --- | --- | --- | --- | --- | --- | --- | --- |
| Altinbilek et al. 2018<sup>7</sup> | Fair | 12 | Y | Y | N | N | Y | Y | Y | Y | Y | Y | Y | Y | Y | Y |
| Schwerla et al. 2020<sup>6</sup> | Fair | 12 | Y | Y | N | N | Y | Y | Y | Y | Y | Y | Y | Y | Y | Y |
| Sezerel et al. 2024<sup>8</sup> | Fair | 12 | Y | Y | N | N | Y | Y | Y | Y | Y | Y | Y | Y | Y | Y |

Three RCTs were included in our statistical analysis. A total of 231 participants were assessed who received structural pain management. Of the 231 participants, 117 patients received OMT to aid in pain management, while 114 patients received pain management in different modalities. The outcomes measured between the three studies in our meta analysis was the intensity of pain on the VAS scale. Secondary outcomes were not measured due to inconsistencies of outcomes observed in the studies. The random effects model was used in evaluating outcomes within the three studies.

Figure 2 details the random-effects model of evaluating pain intensity on the VAS scale following treatment.

**Figure 2:**
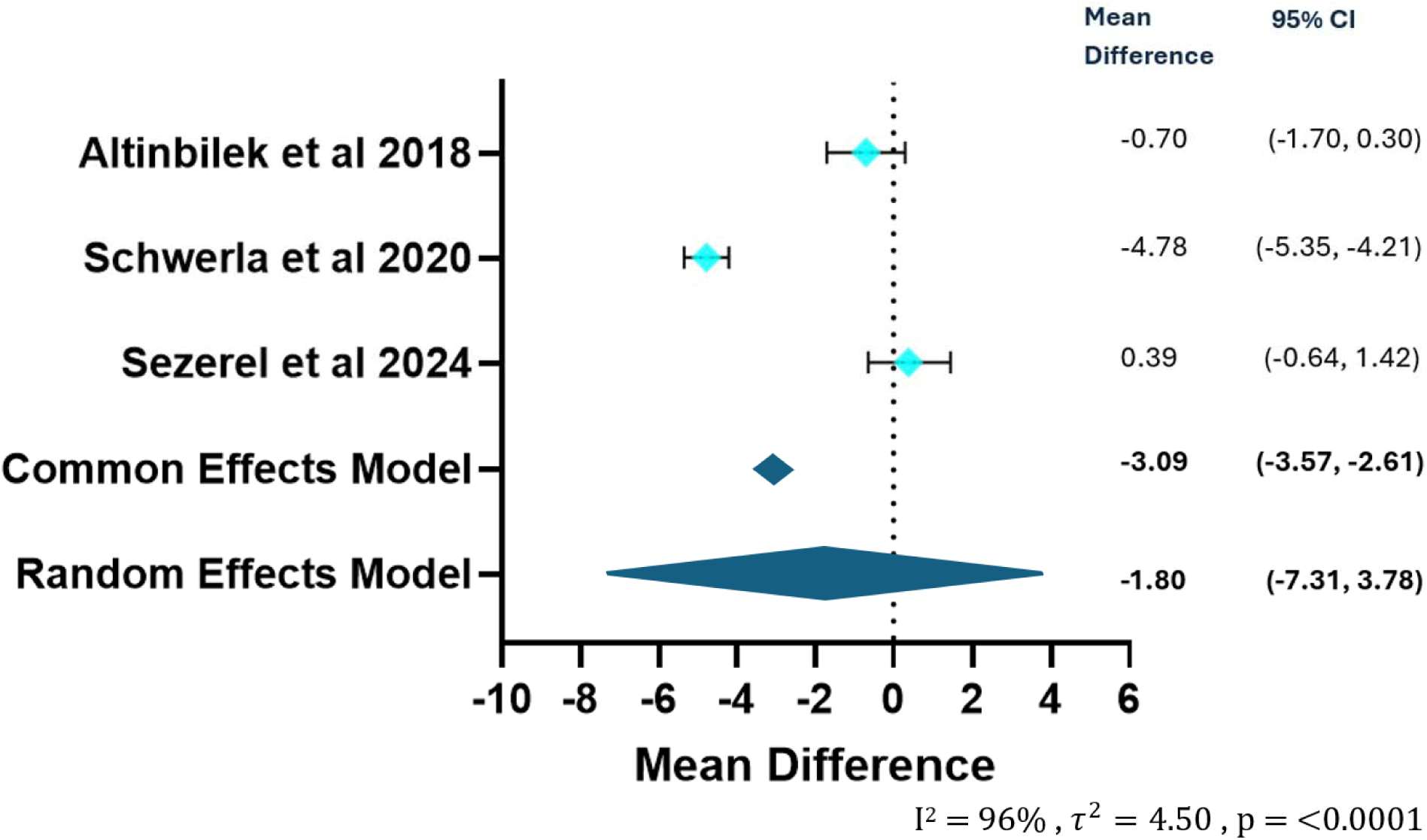
Forest plot of the mean difference in pain reduction for OMT groups and non OMT groups treated for structural pain. CI: confidence interval; OMT: osteopathic manipulative treatment

Figure 2 shows the random-effects model for pain intensity assessed by VAS scale for patients treated with structural pain for Altinbilek et al. 2018^7^, Schwerla et al. 2020^6^, and Sezerel et al. 2024^8^. The mean difference observed was -1.80 (−7.31; 3.78) (Figure 2). The confidence interval includes zero, which indicates that there is no statistically significant difference between pain intensity post treatment in the OMT and non-OMT treated groups (Figure 2). Two of three studies favored lower post treatment pain intensity levels in the OMT group while one study (Sezerel et al^6^) favored lower post treatment pain intensity in the non-OMT treated group. The overall effect sizes between the studies was largely equal. Schwerla et al. 2020^6^ contributed the largest weight to the random effects model at 34.2%, whereas Altinbilek et al. 2018^7^ contributed at 33.0% and Sezerel et al. 2024^8^ contributed the smallest weight at 32.8%. Schwerla et al. 2020^6^ also proved to have the smallest confidence interval (−5.35; -4.21) which indicates that the study had the most precision between the three studies. Heterogeneity was found to be extraordinarily high (I^2^ = 96%) which was found to be statistically significant (p = <0.0001), indicating rejection of the null hypothesis of no heterogeneity (Figure 2).

Figure 3 shows the funnel plot that assesses for publication bias of the included studies evaluating pain intensity on the VAS scale following treatment.

**Figure 3:**
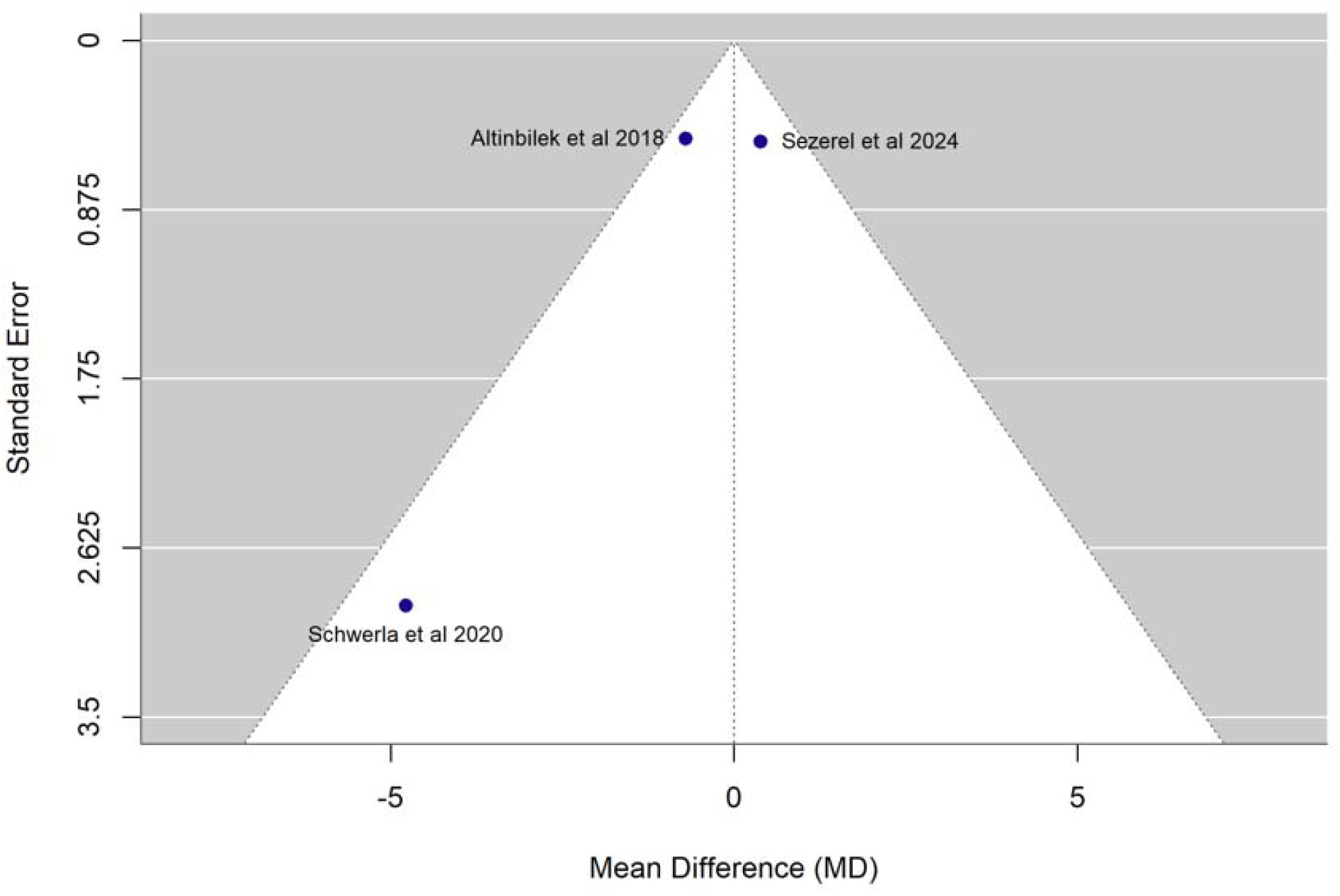
Funnel plot of publication bias in regard to pain reduction of OMT groups and non OMT groups treated for structural pain. OMT: osteopathic manipulative treatment

The funnel plot shown in Figure 3 illustrates assessment of risk of publication bias across included studies evaluating reduction of pain intensity post treatment. A relatively balanced distribution can be observed with effects distributed around the midline. Altinbilek et al. 2018^7^ and Sezerel et al. 2024^8^ appear near the upper portion of the plot with smaller standard errors, reflecting more precise estimates, with Altinbilek et al. 2018^7^ clustering just to the left and Sezerel et al. just to the right of the vertical midline. Schwerla et al. 2020^6^ appears near the lower left portion of the plot with a larger standard error, indicating a less precise estimate given the study’s variance parameters. Overall, no study falls outside of the triangular funnel limits, and the positioning of the points suggests a low likelihood of publication bias for the evaluated clinical outcomes (Figure 3).

Figure 4 details the random-effects model of evaluating pain intensity on the VAS scale following treatment excluding Schwerla et al, the outlier.

**Figure 4:**
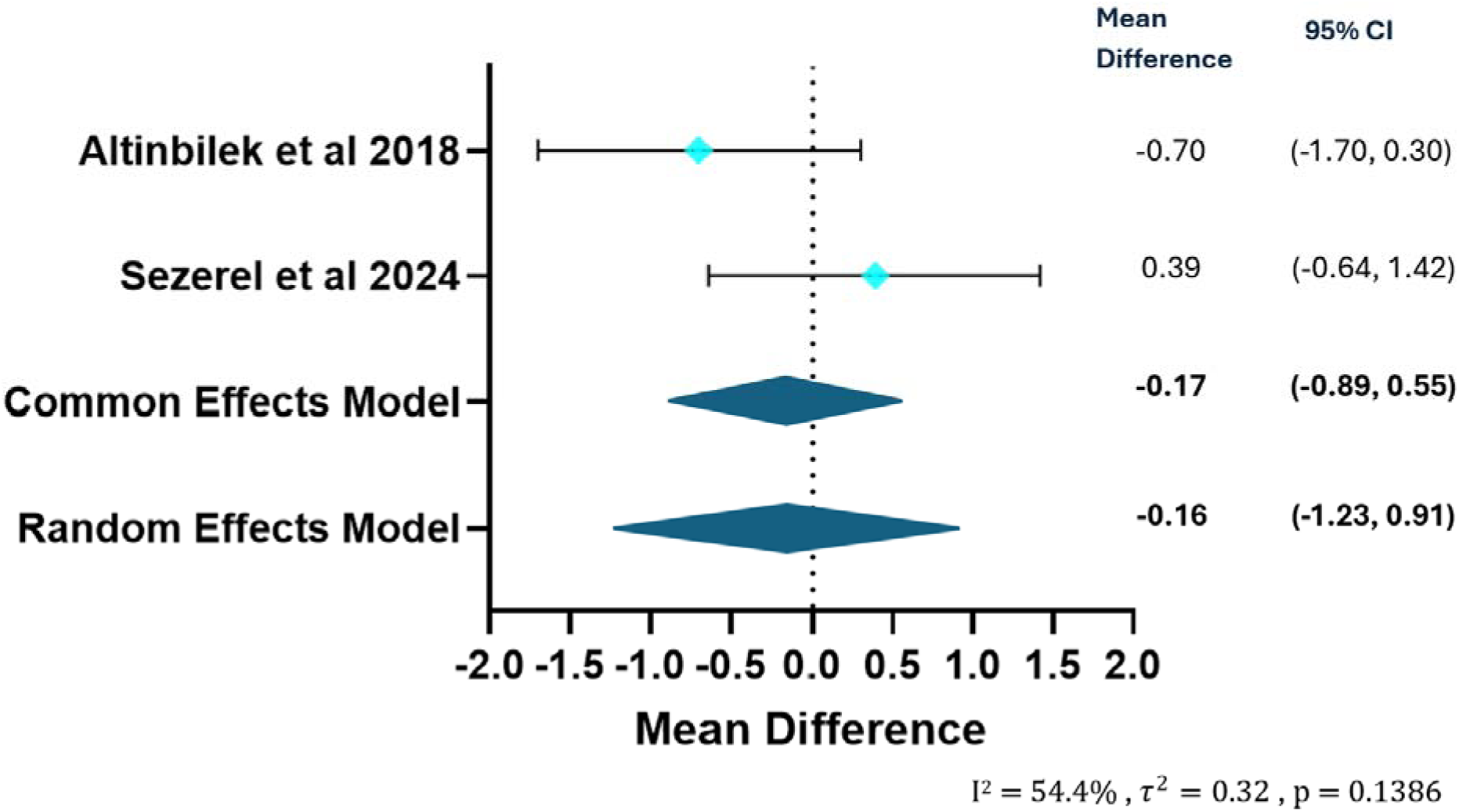
Forest plot of the mean difference in pain reduction for OMT groups and non OMT groups treated for structural pain excluding the outlier (Schwerla et al.). CI: confidence interval; OMT: osteopathic manipulative treatment

Figure 4 shows the random-effects model for pain intensity assessed by VAS scale for patients treated with structural pain for Altinbilek et al. 2018^7^ and Sezerel et al. 2024^8^. The mean difference observed was -0.16 (−1.23; 0.91) (Figure 4). The confidence interval includes zero, which indicates that there is no statistically significant difference between pain intensity post treatment in the OMT and non-OMT treated groups (Figure 4). One study (Altinbilek et al. 2018^7^) favored lower post treatment pain intensity levels in the OMT group while one study (Sezerel et al. 2024^8^) favored lower post treatment pain intensity in the non-OMT treated group. The overall effect sizes between the studies was largely equal. Altinbilek et al. 2018^7^ contributed the largest weight to the random effects model at 50.4%, whereas Schwerla et al. 2020^6^ contributed 49.6%. Altinbilek et al. 2018^7^ also proved to have the smallest confidence interval (− 1.70; 0.30), indicating that the study had more precision than Sezerel et al. 2024^8^ (−0.64; 1.42). Heterogeneity was found to be moderate (I^2^ = 54.4%) which was found to be not statistically significant (p = 0.1386), indicating the null hypothesis of no heterogeneity could not be rejected (Figure 4).

Figure 5 shows the funnel plot that assesses for publication bias of the included studies, excluding the outlier (Schwerla et al. 2020^6^), evaluating pain intensity on the VAS scale following treatment.

**Figure 5:**
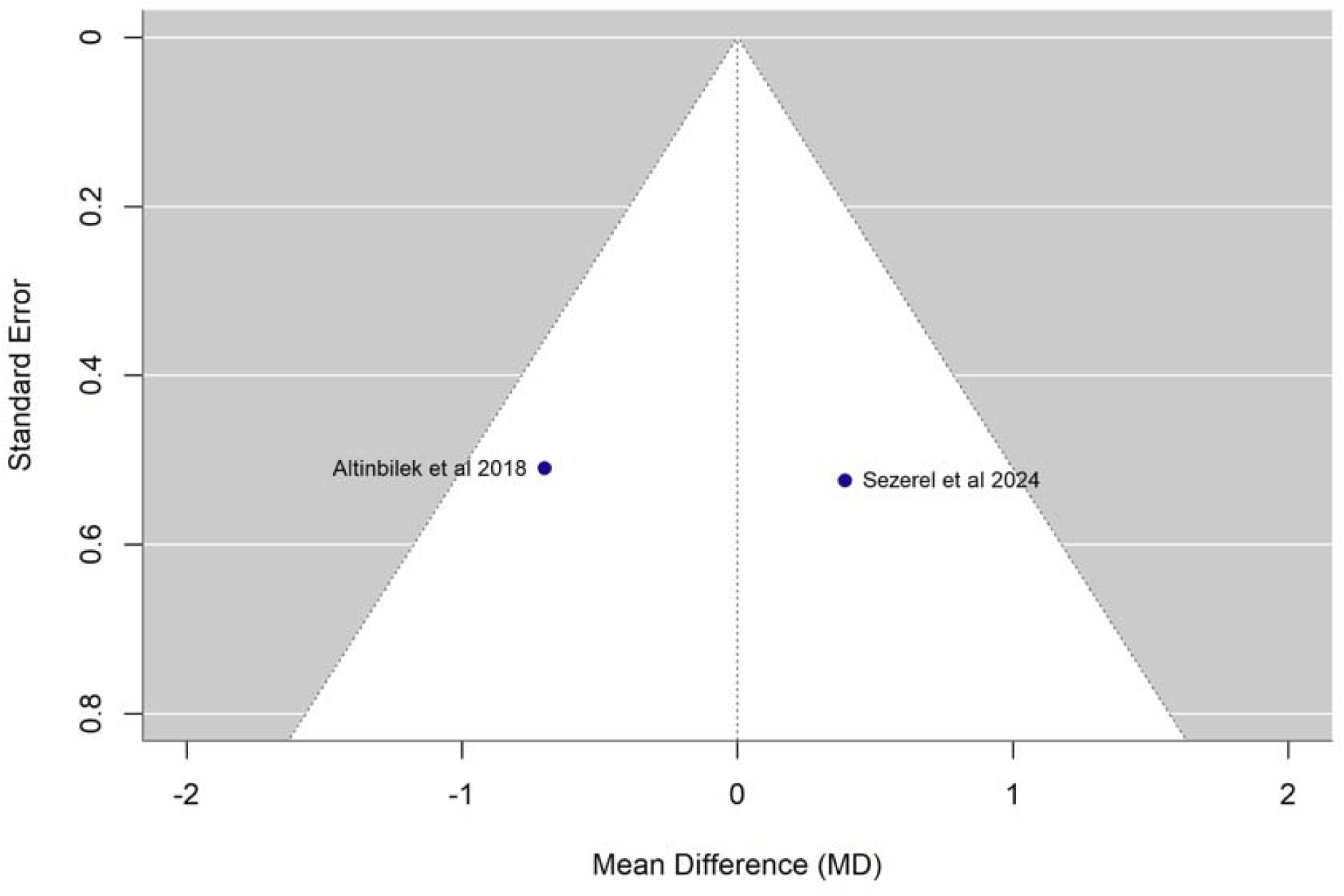
Funnel plot of publication bias in regard to pain reduction of OMT groups and non OMT groups treated for structural pain excluding the outlier (Schwerla et al.). OMT: osteopathic manipulative treatment

The funnel plot shown in Figure 4 illustrates assessment of risk of publication bias across included studies evaluating reduction of pain intensity post treatment excluding the outlier (Schwerla et al. 2020^6^). A highly symmetrical distribution can be appreciated with the two included studies evenly spaced from the vertical midline. Altinbilek et al. 2018^7^ appears in the middle portion of the plot to the left of the midline, while Sezerel et al. 2024^8^ mirrors this positioning to the right of the midline. Both studies cluster at a similar depth on the vertical axis (Standard Error 0.51 and 0.52 respectively), reflecting nearly identical sample sizes and precision parameters. Overall, neither study falls outside of the triangular funnel limits, and the balanced, mirrored positioning of the points suggests a low likelihood of publication bias within this specific subgroup (Figure 4).

## Discussion

The objective of this meta-analysis was to understand and evaluate the effectiveness of osteopathic manipulative treatment (OMT) as an adjunct treatment compared to conventional treatments alone in reducing structural pain. We hypothesized that care containing OMT would result in a greater reduction in structural pain than treatments without it. Data was collected from randomized controlled trials and compared to evaluate the efficacy of OMT in the reduction of structural pain.

To evaluate this, papers were screened for OMT use in relation to structural pain with 157 total research articles identified. Of that, 8 were identified as being randomized controlled trials and 3 were used as being published since 2016.

During our analysis, we examined the heterogeneity (I squared) of the papers which was at 96% due to an outlier. This indicates that there were real differences in the clinical setups of the studies as opposed to the effect being due to random chance. With removal, we found that the heterogeneity would become 54.4% leading to a more uniform examination of the studies. This means there is no “true” effect of OMT on pain, and that effect varies based on how the study is performed. With studies such as Schwela et al. 2020^6^ we see that OMT as a standalone treatment can make a significant impact on pain. Contrarily, OMT used in conjunction with other forms of pain management such as an exercise plan in Altinbilek et al. 2018^7^ or Sezerel et al. 2024^8^, shows the effect of OMT becomes diluted.

Despite this, there were several limitations to the study with the most prominent being the sample size. While 8 papers met the eligibility criteria of being randomized controlled trials, it was restricted to only 3 papers based on publication date. This limit in the current literature reduces the statistical power of the analysis. Additionally, the papers evaluated lacked uniformity. Examining the heterogeneity of the papers, it was found to be 96%. With removal of one of the papers, a clear outlier, it drops to 54.4%. Lastly, OMT techniques can vary. Techniques can be performed in different ways based on the provider, leading to variations in treatment and outcomes.

## Conclusion

This meta-analysis found insufficient evidence of a statistically significant difference in pain intensity, measured using the VAS, between OMT-containing care and non-OMT care for adults with structural musculoskeletal pain. Two of the three studies included confidence intervals that crossed zero, indicating that their findings were not statistically significant, while one study demonstrated a statistically significant benefit favoring OMT. Overall, the findings were not consistent enough to support our hypothesis that OMT-containing care would result in greater structural musculoskeletal pain reduction.

Despite the lack of consistent statistical significance, modest improvements in pain may still be meaningful for some patients, particularly when they improve daily activities, mobility, and quality of life. The decision to include OMT in a treatment plan should depend on the patient’s underlying condition and preferences, as well as the physician’s clinical judgment and experience. Additional high-quality randomized controlled trials with larger sample sizes, standardized treatment protocols, and consistent pain measurements are needed to better determine which patients and conditions may benefit from OMT. Further research will help clarify the role of OMT and support more informed, evidence-based decisions regarding its use in structural musculoskeletal pain management.

## Data Availability

All data produced in the present work are contained in the manuscript.

https://www.nhlbi.nih.gov/health-topics/study-quality-assessment-tools.

https://doi.org/10.1136/bmj.l4898

## Acknowledgement

We would like to thank Dr. Sherry Li for her contributions to and support of this project.

## References

[1] Bagagiolo D, Rosa D, Borrelli F. Efficacy and safety of osteopathic manipulative treatment: an overview of systematic reviews. BMJ Open 2022; 12(4): e053468.

[2] El-Tallawy SN, Nalamasu R, Salem GI, LeQuang JA, Pergolizzi JV, Christo PJ. Management of Musculoskeletal Pain: An Update with Emphasis on Chronic Musculoskeletal Pain. Pain Ther 2021; 10(1): 181–209.

[3] Page MJ, McKenzie JE, Bossuyt PM, et al.: The PRISMA 2020 statement: an updated guideline for reporting systematic reviews. BMJ. 2021, 372:n71. 10.1136/bmj.n71.

[4] Sterne JAC, Savović J, Page MJ, Elbers RG, Blencowe NS, Boutron I, Cates CJ, Cheng H-Y, Corbett MS, Eldridge SM, Hernán MA, Hopewell S, Hróbjartsson A, Junqueira DR, Jüni P, Kirkham JJ, Lasserson T, Li T, McAleenan A, Reeves BC, Shepperd S, Shrier I, Stewart LA, Tilling K, White IR, Whiting PF, Higgins JPT. RoB 2: a revised tool for assessing risk of bias in randomised trials. BMJ 2019; 366: l4898.

[5] Study quality assessment tools. (2021). Accessed: July 19, 2026: https://www.nhlbi.nih.gov/health-topics/study-quality-assessment-tools.

[6] Schwerla F, Hinse FX, Klosterkamp M, Schmitt T, Rütz M, Resch KL. Osteopathic treatment of patients with shoulder pain. A pragmatic randomized controlled trial. J Bodyw Mov Ther 2020; 24(1): 21–28.

[7] Altýnbilek T, Murat S, Yumuşakhuylu Y, İçağasıoğlu A. Osteopathic manipulative treatment improves function and relieves pain in knee osteoarthritis: A single-blind, randomized-controlled trial. Turk J Phys Med Rehabil 2018; 64(2): 114–120.

[8] Sezerel B, Yüksel İ. Efficacy Comparison of Osteopathic Muscle Energy Techniques and Cervical Mobilization on Pain, Disability, and Proprioception in Cervical Spondylosis Patients. Med Sci Monit 2024; 30: e945149.

